# Intravenous Inotropes and Mortality in Takotsubo Syndrome A Nationwide Cohort Study from the SWEDEHEART Registry

**DOI:** 10.64898/2026.11.19.26368542

**Authors:** Thorsteinn Gudmundsson, Sandeep Jha, Petur Petursson, Truls Råmunddal, Oskar Angerås, Araz Rawshani, Ole Fröbert, Joakim Alfredsson, Christina Ekenbäck, Loghman Henareh, Kristofer Skoglund, Charlotta Ljungman, Entela Bollano, Moman A. Mohammad, Henrik Hagström, Tomas Jernberg, David Erlinge, Björn Redfors, Elmir Omerovic

## Abstract

**Background:** Takotsubo syndrome, which predominantly affects women, is thought to be precipitated by catecholamine excess, yet unstable patients often receive intravenous inotropes. We examined the association between intravenous inotropes and 30-day mortality in a nationwide cohort.

**Methods:** We identified all patients with a first angiographic diagnosis of Takotsubo syndrome in the Swedish Coronary Angiography and Angioplasty Registry from 2008 through August 2025, linked to coronary care and population registers. The primary analysis was a Cox model for 30-day death among patients with a coronary care record, adjusted for age, sex, diabetes, smoking, hypertension, hyperlipidemia, and Killip class, with multiple imputation. Sensitivity analyses varied the covariates and standard errors and examined days 0 to 7 and 8 to 30 separately, and the association was estimated in women and men with an interaction term.

**Results:** Of 3,458 patients (79.4% women), 141 (4.1%) received inotropes: 3.6% of women and 5.8% of men. Thirty-day mortality was 19.9% in treated and 1.7% in untreated patients. The unadjusted hazard ratio was 12.89 (95% CI, 8.20 to 20.26) and the adjusted hazard ratio 6.49 (3.49 to 12.08; P<0.001; E-value, 12.5). In sensitivity analyses, the hazard ratio ranged from 3.37 (1.88 to 6.03), with presenting symptom and laboratory values added, to 6.62 (3.63 to 12.08). The hazard ratio was 8.82 (4.47 to 17.39) for days 0 to 7 and 2.47 (0.65 to 9.36) for days 8 to 30, and 6.43 (3.15 to 13.16) in women and 6.59 (2.79 to 15.58) in men (P=0.96 for interaction).

**Conclusions:** Intravenous inotropes were associated with higher 30-day mortality in Takotsubo syndrome, in both women and men, with most of the excess in the first week.

**Clinical Perspective:** *What Is New?:*

- In a nationwide cohort of 3,458 patients with Takotsubo syndrome, 79% of them women, intravenous inotropes were associated with a sixfold higher adjusted hazard of death within 30 days, most of it in the first week.
- The association was similar in women and men and persisted in patients who were stable on arrival and after exclusion of patients resuscitated before hospital.

*What Are the Clinical Implications?:*

- Together with experimental evidence of catecholamine-mediated injury, these findings support the Heart Failure Association position that catecholaminergic inotropes should generally be avoided in Takotsubo syndrome.
- When hemodynamic support is needed, mechanical circulatory support should be considered, and a randomized trial of support strategies is warranted.

## Introduction

Takotsubo syndrome is an acute heart-failure syndrome that predominantly affects women: nearly 8 of 10 patients are women, most of them postmenopausal.^1,2^ It mimics acute myocardial infarction, is typically triggered by emotional or physical stress,^1–4^ and was initially regarded as benign because left ventricular function usually recovers.^1^ It is not benign: severe in-hospital complications, including cardiogenic shock and death, are as frequent as in acute coronary syndromes, and short-and long-term mortality are similar to those after myocardial infarction.^2,5,6^ Men, who make up about 2 in 10 patients, more often have a physical trigger and have higher rates of shock and in-hospital death.^7^

The syndrome also shows how cardiovascular disease in women has been misunderstood. It was first described in Japan in 1990 and was introduced to the international literature as a novel syndrome in 2001,^1,8^ and its neurohumoral basis was described in 2005.^9^ Because it presents with chest pain or dyspnea, electrocardiographic changes, troponin release, and acute left ventricular dysfunction,^1,3,8^ women who had it before it was recognized were diagnosed and treated as having myocardial infarction, myocarditis, or acute heart failure. More than three decades ago, Healy described the “Yentl syndrome,” in which heart disease in women is taken seriously only when it resembles heart disease in men;^10^ cardiovascular disease in women remains understudied, underrecognized, underdiagnosed, and undertreated.^11^ Takotsubo syndrome shows another side of this problem: because it resembles acute coronary syndromes and heart failure, whose treatments were developed in trials that enrolled mostly men,^12^ women with a different disease have been treated as if they had those conditions.

Women with Takotsubo syndrome may still be harmed by treatment borrowed from other conditions. Randomized evidence on hemodynamic support does not exist,^3,4^ and management has been extrapolated from heart failure and cardiogenic shock. For these conditions, guidelines recommend intravenous inotropes to maintain systemic perfusion,^13^ a recommendation derived from heart-failure research in which women are underrepresented relative to their share of the disease.^12^ In Takotsubo syndrome this extrapolation may be particularly harmful, because catecholamines are central to its pathophysiology. Supraphysiological catecholamine levels are found in the acute phase,^9,14^ adrenergic stimulation reproduces the apical ballooning phenotype in animals,^15–17^ and exogenous catecholamines, including dobutamine given during stress testing, have precipitated the syndrome in patients.^18^ The predominance of postmenopausal women may reflect the loss of estrogen’s modulation of this response: in ovariectomized rats, estrogen supplementation attenuates stress-induced left ventricular dysfunction.^19^ Catecholaminergic inotropes are therefore biologically contradictory in this setting and may worsen dynamic left ventricular outflow-tract obstruction, which is present in roughly a quarter of patients.^20,21^

Consensus documents advise against catecholamines and favor mechanical circulatory support when support is needed,^3,4^ but because the syndrome cannot be distinguished from an acute coronary syndrome before angiography, inotropes continue to be used. We examined the association between intravenous inotropes and 30-day mortality in the nationwide Swedish Web-system for Enhancement and Development of Evidence-based care in Heart disease Evaluated According to Recommended Therapies (SWEDEHEART) registry from 2008 through 2025, including the timing of this association, its dependence on adjustment for presenting severity, and whether it differed between women and men.

## Methods

### Data Availability

The data cannot be shared publicly for ethical and privacy reasons. Data from SWEDEHEART are available for research on application to the SWEDEHEART steering group, subject to ethical approval.

### Study Design and Oversight

This nationwide cohort study used data from SWEDEHEART.^22^ The Swedish Coronary Angiography and Angioplasty Registry (SCAAR) records all coronary angiographies and percutaneous coronary interventions in Sweden, entered by the interventional cardiologist; the Register of Information and Knowledge about Swedish Heart Intensive Care Admissions (RIKS-HIA) records all coronary care unit admissions, including presentation, laboratory values, medical history, drugs on admission, treatments during the stay, and complications. Both are validated against patient records.^22^ Vital status was obtained from the Population Register; the most recent recorded death, on August 13, 2025, defined the end of follow-up. The study was approved by the Regional Ethical Review Board in Gothenburg (Dnr 759-13) and the Swedish Ethical Review Authority (Dnr 2022-03103). Individual consent is not required for registry research in Sweden. Drs Omerovic, Gudmundsson, and Redfors had full access to all the data in the study and take responsibility for its integrity and the data analysis.

### Patients

Takotsubo syndrome has been recorded in SCAAR since 2008 and systematically since 2009 according to registry criteria consistent with the Heart Failure Association position statement^3,23^. We included all patients whose first angiography was coded as occurring between February 2008 and August 2025; the index date was the date of that angiography. The SCAAR record was linked to the RIKS-HIA admission during which the angiography was performed (admission no later than 1 day after, and discharge no earlier than 1 day before, the angiography). Patients whose angiography fell on or after the end of follow-up were excluded. The primary cohort comprised patients with an admission in which the inotrope item was recorded.

### Exposure and Outcome

The exposure was intravenous inotropic treatment at any time during the index hospitalization, recorded in RIKS-HIA. The item does not distinguish between agents (dopamine, dobutamine, norepinephrine, epinephrine, milrinone, and levosimendan); the dose, duration, and time of initiation are not recorded. The outcome was death from any cause within 30 days after the index angiography.

### Statistical Analysis

The primary analysis was a Cox proportional-hazards model adjusted for age, sex, diabetes, smoking status, treated hypertension, treated hyperlipidemia, and Killip class, all recorded in SCAAR at the index procedure (Killip class at arrival in the catheterization laboratory, which was on the day of admission or the next in 78% of patients).^23^ Missing covariates (8.6% for smoking and 8.4% for Killip class) were multiply imputed with chained random forests (20 data sets), and estimates were combined with Rubin’s rules.^24–26^ Proportionality of hazards was assessed with Schoenfeld residuals.

Sensitivity analyses of the primary model entered smoking and Killip class as categorical variables, used standard errors clustered by hospital, and added presenting symptom (the symptom that led the patient to seek care), hemoglobin, and creatinine (the first values recorded during the admission) from RIKS-HIA. The time course was examined in days 0 to 7 and days 8 to 30. Inotrope use was compared between women and men with logistic regression, and the association of inotropes with death with an interaction between inotropes and sex in the primary model and in the model with presenting symptom and laboratory values. The primary model was also fitted separately in women and in men, and the propensity-score analysis included an interaction between inotropes and sex.

Propensity-score analyses used overlap weighting^27^ on a score estimated from 22 baseline variables, including shock and resuscitation before arrival, Killip class, heart rate, blood pressure, electrocardiographic findings, renal function, and hemoglobin. The weighted analysis was also performed in patients who were stable on arrival (without cardiogenic shock, resuscitation before hospital, or Killip class IV) and in those with shock, resuscitation, or Killip class II to IV on arrival, and with period-specific hazard ratios for days 0 to 7 and 8 to 30. It was also repeated after exclusion of patients resuscitated before hospital and with low-molecular-weight heparin and fondaparinux as negative-control exposures, and the primary model and the weighted analysis were also performed in all patients, with those without a coronary care record counted as untreated. For the primary model and the model with presenting symptom and laboratory values, E-values^28^ (the minimum strength of association, on the risk-ratio scale, that an unmeasured confounder would need with both treatment and death to explain the estimate away) were calculated for the point estimate and for the confidence limit closer to 1, treating hazard ratios as risk ratios because 30-day mortality was below 3%. A quantitative bias analysis^29^ examined misclassification of the diagnosis. Assuming that some patients did not have Takotsubo syndrome (positive predictive value of the registry diagnosis, 60% to 95%) and that, in these patients, inotropes carried no excess hazard or the hazard ratio reported for cardiogenic shock of any cause (1.72),^30^ the hazard ratio in Takotsubo syndrome was derived from the observed one as a mixture on the hazard scale, an approximation that was checked by simulation, the hazard ratio in misdiagnosed patients that would make the association null was calculated, and the models were refitted after exclusion of patients with obstructive coronary disease, a discharge diagnosis of myocardial infarction or myocarditis, or both (Supplemental Methods). Confidence intervals were not adjusted for multiplicity and should not be used to infer definitive effects. Analyses were performed with R software, version 4.6.1.

## Results

### Patients

Of 5,340 patients with a first angiographic diagnosis of Takotsubo syndrome, 11 had no follow-up time and were excluded; 3,458 (65%) had a coronary care record with the inotrope item recorded and formed the primary cohort; the other 1,871 had no coronary care record with the inotrope item. The 3,458 patients were admitted to 72 hospitals. Intravenous inotropes were given to 141 patients (4.1%). Age, diabetes, smoking, hypertension, and hyperlipidemia differed little between the groups, but treated patients were sicker (Table 1 and Table S1): 45% were in Killip class II to IV (versus 8%), 16% had been resuscitated before hospital (versus 1%), 12% were in cardiogenic shock on arrival (versus 0.5%), and during the stay cardiogenic shock was recorded in 28% (versus 0.6%), resuscitated cardiac arrest in 18% (versus 1.4%), and noninvasive ventilation in 40% (versus 3.8%). The median length of stay was 10 days in treated and 3 days in untreated patients.

**Table 1.** Characteristics of the Patients at Baseline and during the Index Hospitalization.*

| Characteristic | Inotropes (N=141) | No inotropes (N=3,317) | Standardized difference<sup>†</sup> |
|---|---|---|---|
| **Age (yr)** | 71 (62–77) | 70 (62–77) | 0.04 |
| **Female sex** | 100 (70.9) | 2,646 (79.8) | −0.21 |
| **Diabetes** | 24 (17.8) | 415 (12.6) | 0.14 |
| **Current smoker** | 22 (21.6) | 485 (15.9) | 0.15 |
| **Treated hypertension** | 72 (55.8) | 1,679 (51.3) | 0.09 |
| **Treated hyperlipidemia** | 35 (27.1) | 957 (29.4) | −0.05 |
| **Previous myocardial infarction** | 22 (15.8) | 391 (11.9) | 0.12 |
| **Previous PCI** | 13 (9.5) | 236 (7.2) | 0.08 |
| **Killip class at angiography** |  |  |  |
| I | 73 (55.3) | 2,802 (92.4) | −0.93 |
| II | 19 (14.4) | 176 (5.8) | 0.29 |
| III | 22 (16.7) | 38 (1.3) | 0.56 |
| IV | 18 (13.6) | 18 (0.6) | 0.52 |
| **Indication for angiography** |  |  |  |
| Non–ST-elevation acute coronary syndrome | 44 (31.2) | 1,927 (58.1) | −0.56 |
| ST-elevation myocardial infarction | 45 (31.9) | 890 (26.8) | 0.11 |
| Cardiac arrest | 23 (16.3) | 24 (0.7) | 0.58 |
| Heart failure or cardiomyopathy | 20 (14.2) | 118 (3.6) | 0.38 |
| Other | 9 (6.4) | 358 (10.8) | −0.16 |
| **Obstructive coronary artery disease**<sup>‡</sup> | 47 (33.8) | 712 (21.6) | 0.27 |
| **Cardiogenic shock on arrival** | 16 (11.7) | 18 (0.5) | 0.48 |
| **Resuscitation before hospital** | 23 (16.5) | 28 (0.9) | 0.58 |
| **Heart rate (beats/min)** | 102 (81–120) | 83 (70–100) | 0.65 |
| **Systolic blood pressure (mmHg)** | 130 (106–158) | 146 (129–166) | −0.40 |
| **ST elevation** | 49 (35.5) | 989 (30.3) | 0.11 |
| **Creatinine (µmol/L)** | 82 (61–106) | 68 (59–82) | 0.32 |
| **Hemoglobin (g/L)** | 133 (118–146) | 136 (127–145) | −0.23 |
| **Troponin T (ng/L)**<sup>§</sup> | 846 (445–2,077) | 366 (130–750) | 0.49 |
| **Cardiogenic shock during stay**<sup>¶</sup> | 40 (28.4) | 20 (0.6) | 0.86 |
| **Resuscitated cardiac arrest during stay** | 25 (17.7) | 46 (1.4) | 0.58 |
| **Noninvasive ventilation** | 55 (39.6) | 125 (3.8) | 0.96 |
| **Balloon pump or other mechanical support** | 3 (2.1) | 0 | 0.21 |
| **Left ventricular ejection fraction <40%** | 71 (52.6) | 747 (25.7) | 0.57 |
| **Intravenous diuretics** | 112 (79.4) | 502 (15.1) | 1.68 |
| **Length of stay (days)** | 10 (5–14) | 3 (2–5) | 0.77 |
| **Death in hospital** | 26 (18.6) | 38 (1.2) | 0.61 |
| **Death within 30 days** | 28 (19.9) | 57 (1.7) | 0.61 |

<sup>\*</sup>Continuous variables are medians (interquartile ranges); categorical variables are numbers of patients (percentages). Percentages are of patients with the variable recorded; missing values were below 2% except for smoking (8.6%), Killip class (8.4%), creatinine (6.4%), hemoglobin (11.4%), troponin T (38.7%), and ejection fraction (11.9%). Age, sex, diabetes, smoking, hypertension, hyperlipidemia, Killip class, indication, and coronary findings are from SCAAR; the remaining variables are from RIKS-HIA, except balloon pump or other mechanical support (both registries) and deaths (Population Register). The full table with all characteristics is Table S1. PCI denotes percutaneous coronary intervention; RIKS-HIA, Register of Information and Knowledge about Swedish Heart Intensive Care Admissions; SCAAR, Swedish Coronary Angiography and Angioplasty Registry.
<sup>†</sup>The standardized difference is the difference in means or proportions divided by the pooled standard deviation; values above 0.1 in absolute terms indicate imbalance.
<sup>‡</sup>Stenosis of 50% or more in any coronary artery (one-vessel or multivessel disease).
<sup>§</sup>Highest value during the admission.
<sup>¶</sup>Includes shock present on arrival. In the registry's definition, cardiogenic shock includes the need for intravenous inotropes or a balloon pump to maintain blood pressure.

### Primary Analysis

Within 30 days, 28 of 141 treated patients (Kaplan–Meier estimate, 19.9%; 95% CI, 13.0 to 26.2) and 57 of 3,317 untreated patients (1.7%; 95% CI, 1.3 to 2.2) died (Figure 1). The curves separated within days: by day 7, mortality was 17.0% and 1.1%, and 60 of the 85 deaths had occurred. The unadjusted hazard ratio was 12.89 (95% CI, 8.20 to 20.26), and the adjusted hazard ratio in the primary model was 6.49 (95% CI, 3.49 to 12.08; P<0.001), with an E-value of 12.5 (6.4 for the lower confidence limit) (Table 2 and Figure 2). Harrell’s C statistic was 0.83, and there was no evidence of nonproportional hazards for inotropes (P=0.35).

**Figure 1.**
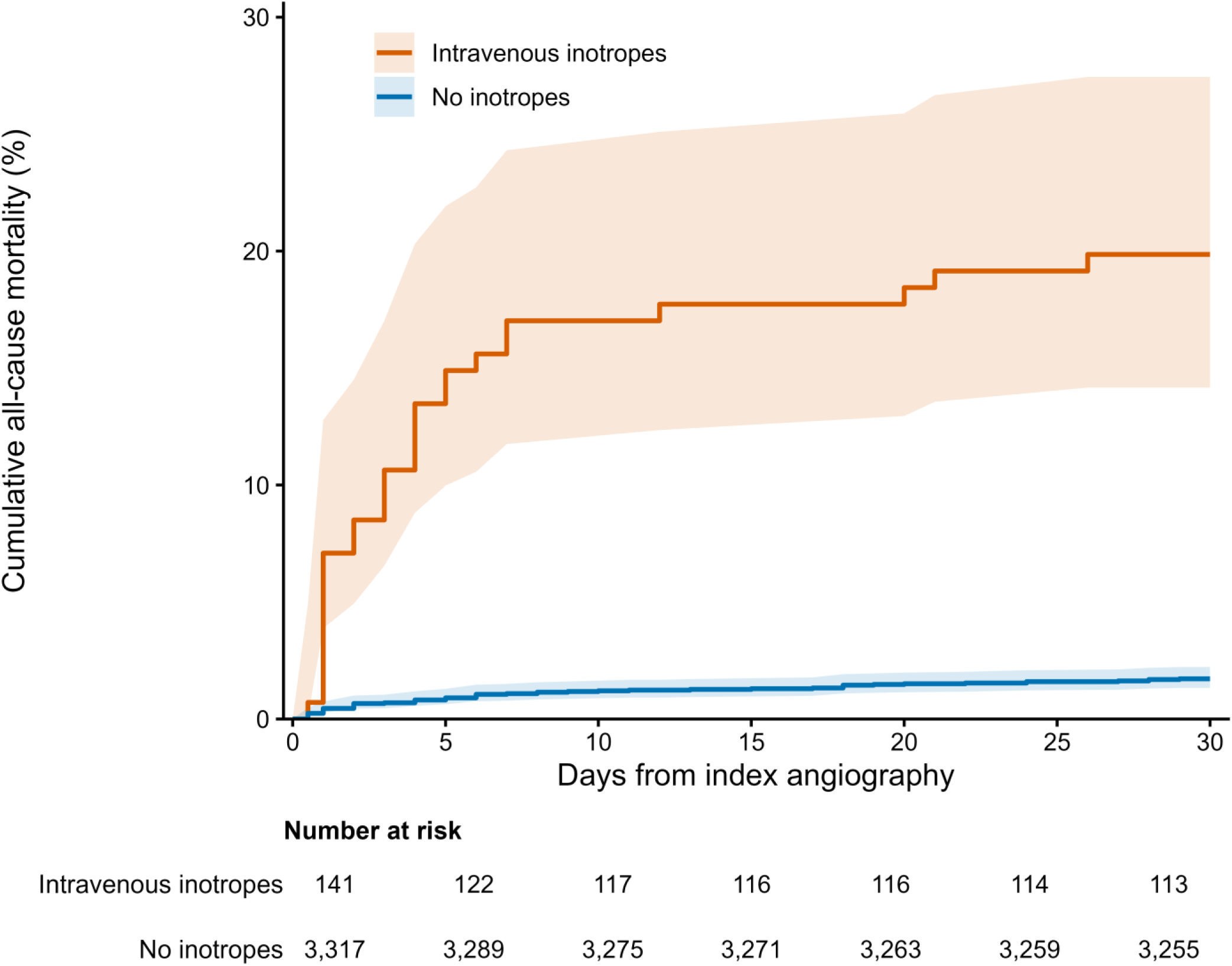
Cumulative mortality within 30 days after the index angiography. Shown are Kaplan–Meier estimates of death from any cause among patients with Takotsubo syndrome who received intravenous inotropes (141 patients) and those who did not (3,317 patients), with pointwise 95% confidence intervals (shaded) and numbers at risk.

**Figure 2.**
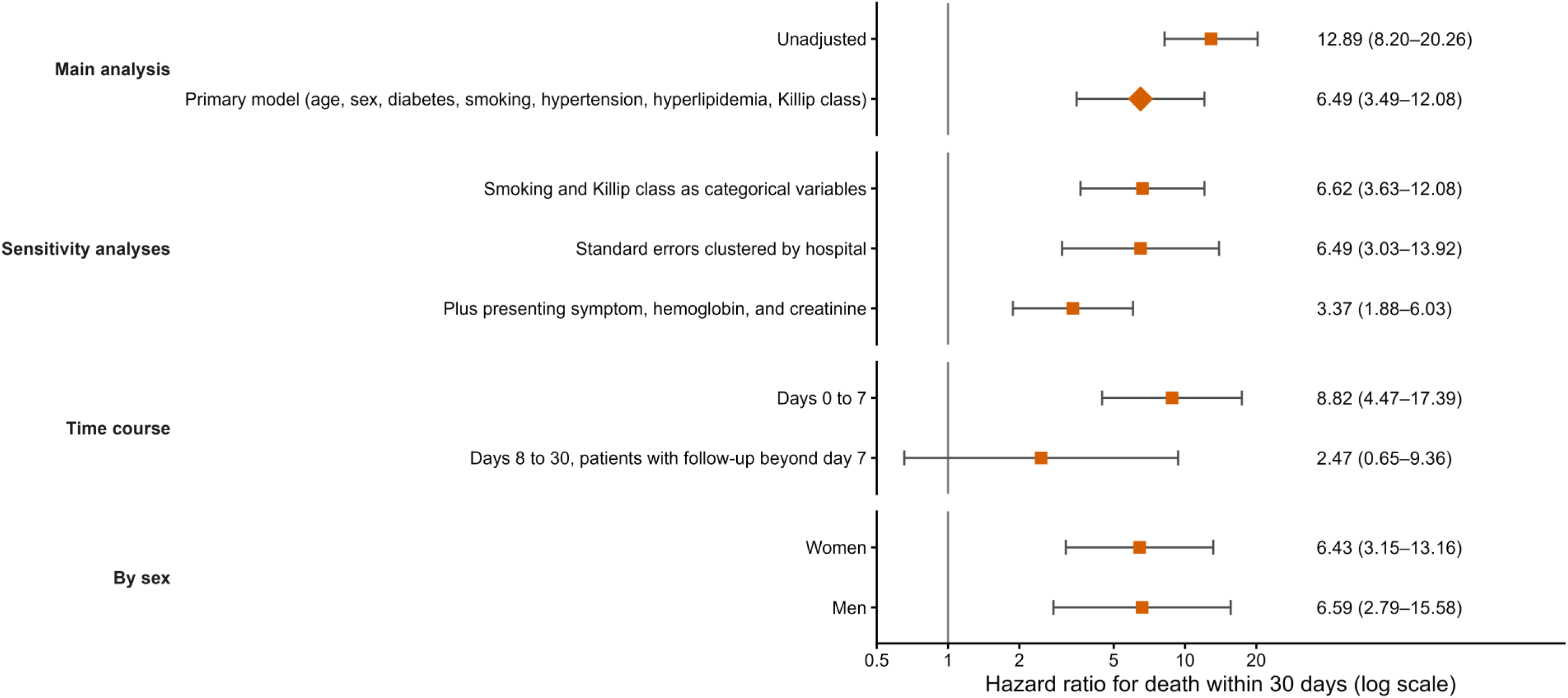
Hazard ratios for death within 30 days associated with intravenous inotropes. Shown are hazard ratios and 95% confidence intervals from the unadjusted model, the primary model, the sensitivity analyses, the analyses by period after the index angiography, and the primary model in women and men, estimated with an interaction between inotropes and sex (Table 2). Confidence intervals were not adjusted for multiplicity.

**Table 2.**
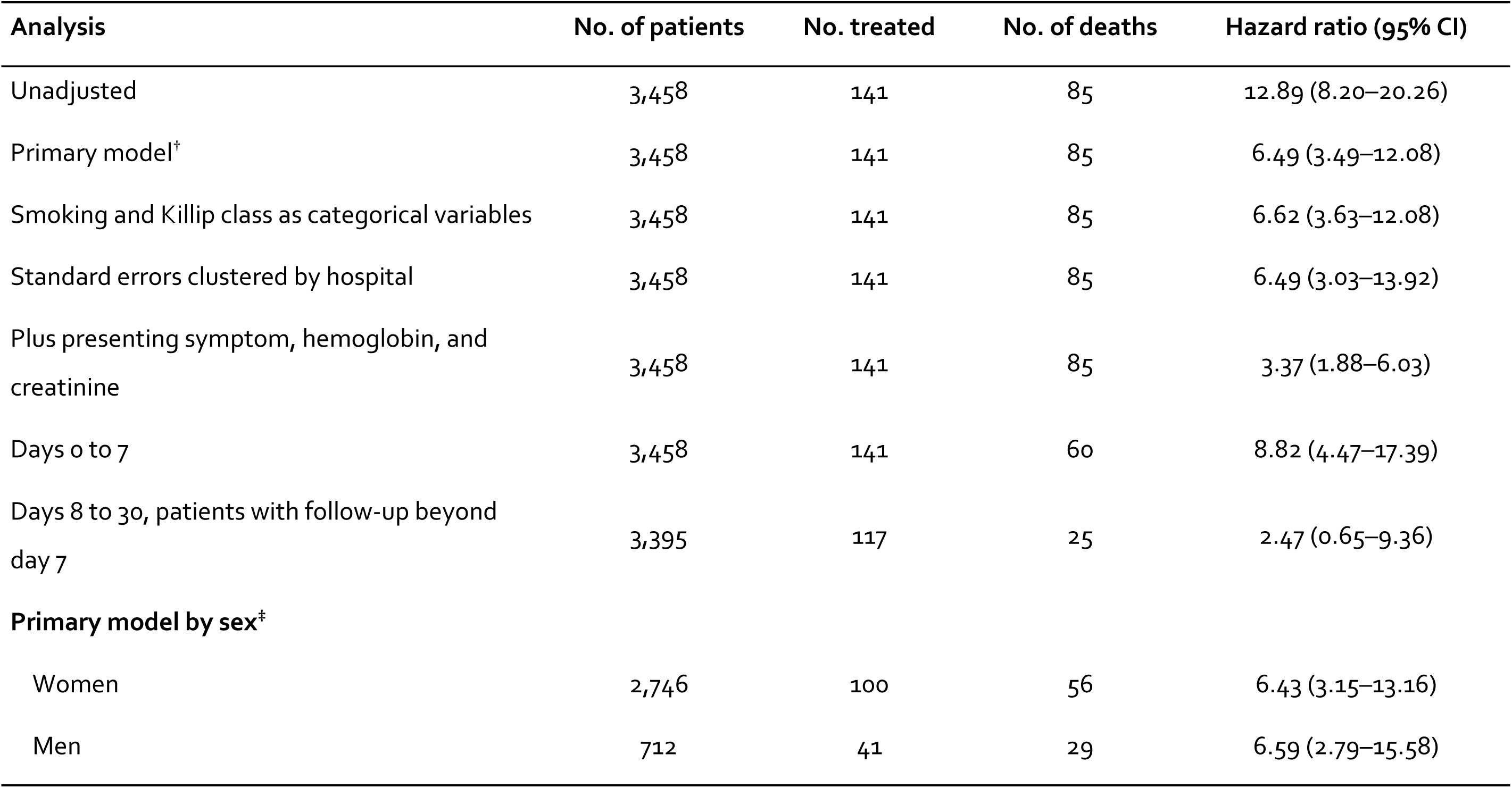
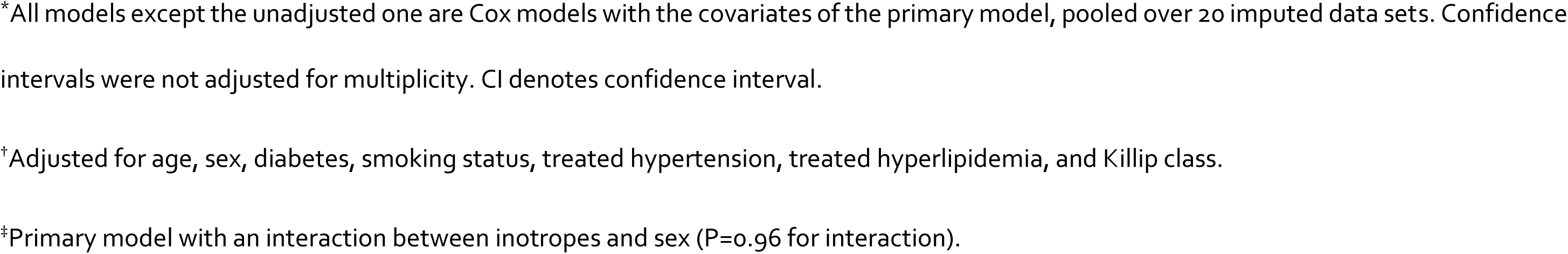
Intravenous Inotropes and Death within 30 Days in the Primary Analysis, Sensitivity Analyses, and Analyses by Sex.*

| **Analysis** | **No. of patients** | **No. treated** | **No. of deaths** | **Hazard ratio (95% CI)** |
|---|---|---|---|---|
| Unadjusted | 3,458 | 141 | 85 | 12.89 (8.20–20.26) |
| Primary model<sup>†</sup> | 3,458 | 141 | 85 | 6.49 (3.49–12.08) |
| Smoking and Killip class as categorical variables | 3,458 | 141 | 85 | 6.62 (3.63–12.08) |
| Standard errors clustered by hospital | 3,458 | 141 | 85 | 6.49 (3.03–13.92) |
| Plus presenting symptom, hemoglobin, and creatinine | 3,458 | 141 | 85 | 3.37 (1.88–6.03) |
| Days 0 to 7 | 3,458 | 141 | 60 | 8.82 (4.47–17.39) |
| Days 8 to 30, patients with follow-up beyond day 7 | 3,395 | 117 | 25 | 2.47 (0.65–9.36) |
| **Primary model by sex<sup>‡</sup>** |  |  |  |  |
| Women | 2,746 | 100 | 56 | 6.43 (3.15–13.16) |
| Men | 712 | 41 | 29 | 6.59 (2.79–15.58) |

<sup>\*</sup>All models except the unadjusted one are Cox models with the covariates of the primary model, pooled over 20 imputed data sets. Confidence intervals were not adjusted for multiplicity. CI denotes confidence interval.
<sup>†</sup>Adjusted for age, sex, diabetes, smoking status, treated hypertension, treated hyperlipidemia, and Killip class.
<sup>‡</sup>Primary model with an interaction between inotropes and sex (P=0.96 for interaction).

### Sensitivity Analyses

The estimate was unchanged with categorical covariates (hazard ratio, 6.62) or hospital-clustered standard errors (6.49; 95% CI, 3.03 to 13.92). Additional adjustment for presenting symptom, hemoglobin, and creatinine reduced it to 3.37 (95% CI, 1.88 to 6.03; E-value, 6.2) (Table 2 and Figure 2). In the overlap-weighted propensity-score analysis, the risk ratio was 3.42 (95% CI, 1.90 to 6.16); it was 7.04 (95% CI, 3.18 to 15.62) among patients who were stable on arrival (95 treated and 3,265 untreated) and 1.89 (95% CI, 0.96 to 3.75) among the 306 patients with shock, resuscitation, or Killip class II to IV on arrival (Table S2). The association persisted after exclusion of patients resuscitated before hospital (risk ratio, 4.10; 95% CI, 2.10 to 8.01) and when the 1,871 patients without a coronary care record were counted as untreated (hazard ratio, 3.57; 95% CI, 1.91 to 6.68; overlap-weighted risk ratio, 2.00; 95% CI, 1.22 to 3.26), whereas neither negative-control drug was associated with death (low-molecular-weight heparin: risk ratio, 0.85; 95% CI, 0.50 to 1.45; fondaparinux: 0.54; 95% CI, 0.28 to 1.07) (Table S2). Correction for misclassification of the diagnosis increased the estimates: had 10% of the patients not had Takotsubo syndrome and inotropes not affected their risk, the hazard ratio among patients with the syndrome would have been 7.10 (95% CI, 3.77 to 13.31) in the primary model and 3.63 (95% CI, 1.98 to 6.59) with fuller adjustment, and had 40% been misdiagnosed, 10.16 (95% CI, 5.15 to 19.46) and 4.94 (95% CI, 2.47 to 9.39). For the association to be null, inotropes would have had to be associated with a hazard ratio of at least 14.7 in misdiagnosed patients (6.9 with fuller adjustment) even if 40% of the patients had been misdiagnosed. Among the 1,971 patients without obstructive coronary disease or a discharge diagnosis of myocardial infarction or myocarditis, the hazard ratio was 11.92 (95% CI, 5.31 to 26.79) in the primary model and 4.99 (95% CI, 1.96 to 12.72) with fuller adjustment (Table S4).

### Time Course

The hazard ratio was 8.82 (95% CI, 4.47 to 17.39) for days 0 to 7 and 2.47 (95% CI, 0.65 to 9.36) for days 8 to 30 (Table 2 and Figure 2); in the overlap-weighted analysis, it was 4.41 (95% CI, 1.87 to 10.41) and 1.18 (95% CI, 0.28 to 4.91), respectively (Table S2).

### Women and Men

Of the 3,458 patients, 2,746 (79.4%) were women. Inotropes were given to 100 women (3.6%) and 41 men (5.8%) (odds ratio, 0.62; 95% CI, 0.43 to 0.90). The difference was smaller after adjustment for the covariates of the primary model (odds ratio, 0.68; 95% CI, 0.44 to 1.05) and, in addition, for presenting symptom, shock on arrival, and resuscitation before hospital (odds ratio, 0.78; 95% CI, 0.49 to 1.24); men had more often been resuscitated before hospital (3.0% versus 1.1%) and were more often in Killip class III or IV (4.3% versus 2.7%). Among patients who were unstable on arrival (cardiogenic shock, resuscitation before hospital, or Killip class IV), 30 of 67 women (44.8%) and 17 of 35 men (48.6%) received inotropes. Within 30 days, 16 of 100 treated women (16.0%) and 40 of 2,646 untreated women (1.5%) died, as did 12 of 41 treated men (29.3%) and 17 of 671 untreated men (2.5%). The adjusted hazard ratio for death was 6.43 (95% CI, 3.15 to 13.16) in women and 6.59 (95% CI, 2.79 to 15.58) in men (P=0.96 for interaction), and with presenting symptom and laboratory values added, 4.26 (95% CI, 2.22 to 8.18) and 2.11 (95% CI, 0.86 to 5.16) (P=0.18 for interaction) (Table 2, Figure 2, and Table S3). Fitted separately in each sex, the primary model gave hazard ratios of 8.12 (95% CI, 3.94 to 16.73) in women and 4.75 (95% CI, 1.77 to 12.76) in men, and in the overlap-weighted analysis the odds ratios were 7.45 (95% CI, 2.95 to 18.80) and 2.91 (95% CI, 0.88 to 9.57) (P=0.28 for interaction) (Table S3).

## Discussion

In this nationwide cohort of 3,458 patients with Takotsubo syndrome admitted to coronary care units over 17 years, intravenous inotropes were associated with 30-day mortality of 20%, as compared with 1.7% among untreated patients, and with a sixfold higher adjusted hazard of death in the primary model. The point estimate exceeded 1 in every Cox model, but its size depended on the covariates: 12.9 unadjusted, 6.5 in the primary model, and 3.4 with presenting symptom and laboratory values added. The association persisted in propensity-score analyses that included presentation severity (risk ratio, 3.4) and was not confined to patients who were unstable on arrival: among those without shock, resuscitation, or Killip class IV on arrival, the risk ratio was 7.0. Only strong unmeasured confounding could explain these associations away: an unmeasured factor would need to be associated with both inotrope use and death by a risk ratio of at least 12.5 (6.4 for the lower confidence limit) beyond the measured covariates, and by at least 6.2 (3.2) for the model with presenting symptom and laboratory values. Misclassification of the diagnosis would have biased the estimates toward the null unless inotropes were more harmful in misdiagnosed patients than in Takotsubo syndrome; even if 40% of the patients had been misdiagnosed, the association would be null only if the hazard ratio in those patients had been at least 15 (7 with fuller adjustment), as compared with 1.7 in cardiogenic shock of any cause.^30^ Consistent with such dilution, the estimates were larger when patients with obstructive coronary disease or a discharge diagnosis of myocardial infarction or myocarditis were excluded. Most of the excess occurred in the first week (hazard ratio, 8.8; 4.4 in the propensity-score analysis); for days 8 to 30 the hazard ratios were 2.5 and 1.2, with confidence intervals that included no effect. The association did not differ between women and men (hazard ratios, 6.4 and 6.6).

These findings are consistent with the central role of catecholamines in the pathophysiology of Takotsubo syndrome.^31,32^ Plasma catecholamine concentrations in the acute phase are markedly higher than in myocardial infarction,^9^ and catecholamines and β-adrenergic agonists given intravenously can precipitate the syndrome immediately, with troponin release, QTc prolongation, and reversible apical, midventricular, or basal ballooning.^33^ Of 157 published cases of drug-induced Takotsubo syndrome, 36% followed the administration of exogenous catecholamines and 68% were related to direct or indirect catecholamine stimulation.^34^ Experimentally, epinephrine, norepinephrine, dopamine, and isoprenaline all induce takotsubo-like dysfunction, in a regional pattern that depends on afterload;^35^ the phosphodiesterase inhibitor milrinone induces apical ballooning on its own, and pretreatment with levosimendan increases its incidence, whereas pretreatment with metoprolol attenuates the dysfunction and reduces mortality.^17^ Because the inotropic effects of β-adrenergic agonists and of phosphodiesterase-3 inhibitors such as milrinone are both mediated by cyclic AMP, the hypothesis of harm extends to most agents recorded under the registry item.

Several mechanisms may make inotropic stimulation particularly harmful once the syndrome has developed. The apical hypokinesia may itself be protective: at high epinephrine concentrations, β2-adrenergic receptors switch from stimulatory to inhibitory G-protein signaling, which depresses apical contraction but protects the cardiomyocytes, and preventing this switch increased mortality experimentally.^15^ Exogenous inotropes can exacerbate left ventricular dysfunction and impede recovery,^31,33^ and catecholamines disturb cardiomyocyte calcium handling, energetics, and electrophysiology^32^ and cause lipotoxic metabolic stunning.^36^ Inotropic stimulation can also aggravate dynamic outflow-tract obstruction, in which positive inotropy lowers blood pressure further,^20,21,37^ and increases oxygen demand and arrhythmia risk in a myocardium with prolonged repolarization.^14,32,33^ Inotropes may likewise induce Takotsubo syndrome in other critically ill patients.^38^ The concentration of the excess deaths in the first week in our cohort is consistent with such a direct drug effect.

In our earlier SWEDEHEART analysis of 1,724 patients treated from 2009 through 2016, a period included in the present cohort, intravenous inotropes were the drug class most strongly associated with death within 30 days (hazard ratio, 9.92).^23^ In the International Takotsubo Registry, catecholamine use was a strong independent predictor of in-hospital death (odds ratio, 9.66; 95% CI, 3.74 to 24.96), and in-hospital mortality with and without catecholamines was 20.6% and 1.6% among women and 27.0% and 2.2% among men.^37^ In our cohort, the corresponding 30-day mortality was 16.0% and 1.5% among women and 29.3% and 2.5% among men. In a German single-center cohort, positive inotropic agents were independently associated with mortality (hazard ratio, 2.2; 95% CI, 1.0 to 4.8),^39^ and among 16,214 Swedish patients with cardiogenic shock of any cause, inotropes were associated with higher 30-day mortality (hazard ratio, 1.72; 95% CI, 1.26 to 2.35).^30^ The Heart Failure Association position statement regards inotropes, including dobutamine, dopamine, norepinephrine, epinephrine, and milrinone, as generally contraindicated in Takotsubo syndrome, because further activation of catecholamine receptors or their downstream pathways might worsen the condition, and suggests mechanical circulatory support, or low-dose levosimendan when this is not available, for cardiogenic shock;^3^ other reviews and consensus documents likewise advise against catecholamines.^4,31,40^

This study has limitations. Confounding by indication cannot be excluded, as treated patients were sicker. The concentration of deaths in the first week may reflect the severity of illness that prompted treatment as well as a drug effect. Because inotropes are given for hemodynamic deterioration, confounding of the E-values by unrecorded severity cannot be excluded. Cardiogenic shock and cardiac arrest before or on arrival may be causes of inotrope treatment or consequences of inotropes given before admission, and the registry does not record their order. The indication, dose, and timing of inotropes are unknown, and drugs given before arrival are not recorded. The diagnosis was recorded by the angiographer and was not adjudicated. Treatment was entered as a fixed covariate from the time origin, so that pretreatment time in patients started later counts as exposed (biasing toward benefit), whereas rescue treatment shortly before death biases toward harm. The comparison of women and men was post hoc, and estimates in men, of whom 41 were treated, are imprecise.

## Conclusions

In a nationwide cohort, intravenous inotropes were associated with higher 30-day mortality in Takotsubo syndrome, sixfold in the primary model and threefold with fuller adjustment, consistently across analytic approaches, in patients who were stable on arrival, and in both women and men, with most of the excess in the first week. Together with experimental and clinical evidence of catecholamine-mediated injury, these findings support the position of the Heart Failure Association that inotropes acting through catecholamine receptors or their downstream pathways should generally be regarded as contraindicated in Takotsubo syndrome, and that mechanical circulatory support, or low-dose levosimendan where it is unavailable, should be considered when hemodynamic support is needed. A randomized trial of hemodynamic support strategies is warranted.

## Data Availability

The data cannot be shared publicly because of Swedish and EU data-protection legislation. SWEDEHEART data are available to researchers on application to the SWEDEHEART steering group, subject to ethical approval.

## Acknowledgments

We thank the staff of the Swedish coronary care units and catheterization laboratories who enter data into SWEDEHEART. Claude (Anthropic) was used to assist with statistical programming, drafting, and language editing; the authors designed the study, checked all analyses and text, and take full responsibility for the content.

## Supplemental Material

Supplemental Methods; Tables S1–S4.

## Non-standard Abbreviations and Acronyms

CI: confidence interval
RIKS-HIA: Register of Information and Knowledge about Swedish Heart Intensive Care Admissions
SCAAR: Swedish Coronary Angiography and Angioplasty Registry
SWEDEHEART: Swedish Web-system for Enhancement and Development of Evidence-based care in Heart disease Evaluated According to Recommended Therapies

